# An epidemic CC1-MRSA-IV clone yields false negative test results in molecular MRSA identification assays: a note of caution

**DOI:** 10.1101/2020.04.28.20083048

**Authors:** Stefan Monecke, Elisabeth König, Megan R. Earls, Eva Leitner, Elke Müller, Gabriel Wagner, David M. Poitz, Lutz Jatzwauk, Teodora Vremerǎ, Olivia S. Dorneanu, Alexandra Simbeck, Andreas Ambrosch, Ines Zollner-Schwetz, Robert Krause, Werner Ruppitsch, Wulf Schneider-Brachert, David C. Coleman, Ivo Steinmetz, Ralf Ehricht

## Abstract

**Background:** A variety of rapid molecular PCR tests has been developed and commercialised that interrogate the junction site between the staphylococcal core genome, and the mobile genetic element (SCC*mec*) which harbours the gene responsible for methicillin-/beta-lactam-resistance.

**Aim:** The purpose of the present study was to investigate why a clinical MRSA isolate yielded false negative test results with widely used, commercial *orfX*/SCC*mec* junction site PCR tests.

**Methods:** A collection of isolates that belonged to the same epidemic strain as the index isolate were investigated with commercial MRSA assays and all isolates were sequenced in order to explain this observation.

**Results:** It was found that isolates of the epidemic “European CC1-MRSA-IV” clone, which likely originated in South-Eastern Europe and subsequently spread to Western Europe, generally exhibit this behaviour. The failure of the assays was attributable to a characteristic large insertion in the *orfX*/SCC*mec* integration site presumably targeted by such tests. In contrast to MW2 (GenBank BA000033.2, a CC1 “USA400” strain which also harbours SCC*mec* IVa), the European CC1 clone harbours an insertion of *ca*. 5,350 nucleotides adjacent to *orfX*. This sequence starts with a novel SCC terminal sequence alternate to *dcs* and encodes six different hypothetical proteins (E7MHX1, *ydiL*2, C5QAP8, A8YYX4, *npd-*SCC, H4AYD7; nucleotide positions 280,690–286,024 of GenBank RBVO000005.1). An SCC*mec* element with the same insertion was previously found in a *Staphylococcus epidermidis* isolate (GenBank MH188467.1) suggesting transfer between staphylococcal species.

**Conclusion:** In order to ensure the reliability of molecular MRSA tests, it is vital to monitor the emergence of new SCC*mec* junction sites, not only in *Staphylococcus aureus* but also in coagulase-negative staphylococci.

## INTRODUCTION

*Staphylococcus aureus* is a Gram-positive bacterium that frequently colonises both humans and animals, mostly as a harmless commensal of the skin and mucous membranes, especially the nares. However, *S. aureus* can also be pathogenic, causing skin and soft tissue infections, pneumonia, meningitis, endocarditis, mastitis, osteomyelitis, toxic shock syndrome and sepsis. It can harbour numerous virulence-associated genes, many of which reside on mobile genetic elements (MGEs). Infections caused by *S. aureus* can be treated with a variety of antibiotics including flucloxacillin, cefazolin, co-trimoxazole, clindamycin, vancomycin, doxycycline amongst others, depending on the clinical presentation, severity and the susceptibility profile of the infecting strain.

Methicillin-resistant *S. aureus* (MRSA) were first identified almost 60 years ago and now constitute a major obstacle to therapy. Methicillin resistance is mediated by a modified penicillin binding protein (PBP2́ or PBP2a) that confers resistance to virtually all beta-lactam antibiotics, with the exception of some recently developed compounds such as ceftobiprole and ceftaroline. The genes *mecA* or *mecC* typically encode this property. The *mecA* gene is most commonly responsible for methicillin resistance in *S. aureus* and is localised on a variety of large and complex mobile genetic elements (MGEs), called staphylococcal cassette chromosome elements (SCC*me*c). The *mecC* gene is localised on an uncommon SCC*mec* element (type XI; (1, 2)) and its presence is largely restricted to Western Europe where a distinct zoonotic connection to wildlife, especially hedgehogs, has been observed (3–5). Finally, there have been sporadic reports of plasmid-encoded genes, *mecB* (6) and *mecD* (7), which possibly originated in micrococci.

As therapy options for MRSA infections are limited, emphasis must be placed on infection prevention and control (IPC) measures including hand hygiene, the use of protective clothing and equipment and isolation of patients. Effective IPC implementation requires the rapid detection of MRSA. Traditionally, this has been achieved using selective growth media containing methicillin or cefoxitin, salt and a chromogenic substrate (for an overview, see (8)). The latter facilitates the quick differentiation of MRSA from other, clinically less relevant staphylococcal species that may also harbour *mec* genes. While the use of chromogenic media is relatively inexpensive and convenient, plates must be incubated for 18–24 h before results are obtained. Considering that clinical judgements on whether to isolate a patient or to commence presumptive therapy with antibiotics that covers MRSA commonly require a much quicker decision, a variety of rapid molecular PCR tests has been developed and commercialised (9). These need to identify the presence of *mecA* and *mecC* and/or the presence of SCC*mec* elements together with a *S. aureus*-specific genetic marker(s). In addition, molecular tests should remain negative if the *mec* gene and/or the SCC*mec* element are present in another staphylococcal species that also might occur in the same sample. This can be achieved by placing one primer for the amplification reaction into the core chromosomal gene adjacent to the SCC*mec* element (*orfX*), which is present in all staphylococci, the sequence of which differs between different species. Importantly, the second primer cannot be placed into the *mecA* or *mecC* sequence as the distance between *orfX* and the *mec* gene is simply too large to be covered by a standard PCR. A “downstream constant segment” (*dcs*) of SCC is widely distributed and highly conserved among MRSA strains, even among strains harbouring different types of SCC*mec* elements, making it an obvious choice for the placement of a second amplification primer. However, this approach can have drawbacks. Firstly, methicillin-susceptible *S. aureus* might yield a positive test result if they carry an SCC element without a *mec* gene, such as a dysfunctional remnant of SCC*mec* (10) or an element associated with fusidic acid resistance. Secondly, an MRSA strain with a non-*dcs* downstream sequence might yield a negative result in such a test. In a central European setting, ca. 5% of MRSA strains might carry one of as many as fourteen known different SCC terminal sequences alternate to *dcs* (11). In order to ensure the reliability of PCR-based MRSA detection assays, newly obtained sequences of the *orfX*/SCC*mec* junction site should be monitored, and the emergence of new SCC terminal sequences should prompt an update to these tests. Thirdly, there are strains that are phenotypically susceptible to betalactam antibiotics (12) despite being positive in molecular tests targeting *mecA* and/or the *orfX*/SCC*mec* junction site.

The purpose of the present study was to investigate why a clinical isolate yielded false negative test results with a widely used, commercial *orfX*/SCC*mec* junction site PCR tests (Cepheid GeneXpert MRSA/SA BC and BD MAX Staph SR Assay), although it was confirmed to be MRSA by other means. It was found that isolates of the epidemic “European CC1-MRSA-IV” clone, which likely originated in South-Eastern Europe and subsequently spread to Western Europe ((13, 14); for a detailed description, see also below), generally exhibit this behaviour. The failure of the assay was attributed to a characteristic large insertion in the *orfX*/SCC*mec* integration site presumably targeted by this test.

## MATERIALS AND METHODS

### Index case

The index case of the present study was a 62-year-old man with bacteraemia caused by CC1MRSA which yielded a negative result using commercially available MRSA PCR systems in widespread use.

The index patient, who had metastasised lung cancer, was admitted to the Department of Internal Medicine, Medical University of Graz (Austria) with suspected pneumonia, and amoxicillin/clavulanic acid therapy was started. Blood cultures, incubated in an automated BACTEC system (Becton Dickinson; Heidelberg, Germany/Franklin Lakes, NJ, USA) became positive within one day. The organism was identified as *S. aureus* directly from positive blood culture bottles by peptide nucleic acid fluorescence *in situ* hybridisation (PNA FISH, AdvanDx, Woburn, MA, USA). The *S. aureus* isolate from one blood culture bottle (Graz_511421–19) was investigated by using the GeneXpert MRSA/SA BC PCR assay (Cepheid, Sunnyvale, CA, USA) in order to identify *S. aureus*/MRSA. Simultaneously, rapid antimicrobial susceptibility testing (RAST) was performed according to EUCAST guidelines (15, 16). The GeneXpert test was negative for MRSA but RAST revealed cefoxitin resistance after six hours of incubation. Additional antimicrobial susceptibility testing (Vitek2, bioMérieux, Marcy-l’Étoile, France) was performed confirming that it was indeed an MRSA isolate. Another molecular MRSA detection assay, BD MAX^™^ StaphSR assay (Becton Dickinson) was performed; it also yielded a negative result. Microarray-based characterisation with the *S. aureus* Genotyping Kit 2.0 (Abbott [Alere Technologies GmbH], Jena, Germany; see below) detected *mecA*, confirming the disk diffusion result for MRSA, and assigned the isolate to CC1-MRSA-IV.

Initially, before there was evidence for an MRSA infection, the patient́s condition improved under beta-lactam treatment, and therapy was changed to flucloxacillin and fosfomycin. One day later, after antimicrobial susceptibility testing confirmed MRSA with susceptibility to fosfomycin and daptomycin, the treatment was modified and flucloxacillin replaced by daptomycin in addition to fosfomycin. Despite the fact that daptomycin has no pulmonary activity, alternative substances like vancomycin or linezolid were contraindicated due to nephro- and myelotoxicity. Fifth generation cephalosporins were not available. The patient died a few days later because of tumour progression.

The index isolate was identified as European CC1-MRSA-IV using DNA microarray profiling and whole-genome sequencing (see below).

#### Additional CC1-MRSA-IV isolates investigated

Beside the index case, another 38 additional isolates of the European CC1-MRSA-IV clone were investigated.

Ten Romanian CC1-MRSA-IV isolates were recovered between 2007 and 2010, during routine microbiological diagnostics in the Sfanta Parascheva Hospital for Infectious Diseases in Iasi, a 300-bed tertiary centre in North-Eastern Romania, where European CC1-MRSA-IV appears to be common (17).

Four isolates were recovered from diagnostic or screening procedures at the Medical Faculty of Dresden, a 1,200-bed tertiary care hospital in South-Eastern Germany. In Dresden, European CC1-MRSA-IV is very rare, with only six isolates being identified at the University Hospital among 1,752 isolates characterised by microarray profiling between 2000 and 2020 ((11) and unpublished data). In addition to European CC1-MRSA-IV isolates, four control strains were included in this study (see Table 1). Controls were assigned by microarray profiling (see below) to epidemic strains that are or were prevalent locally, or that belonged to another CC1-MRSA strain (isolate #220663). In addition, fully sequenced reference strains MU50, MW2, N315 and USA300-FPR3757 were also included as controls.

**Table 1:**
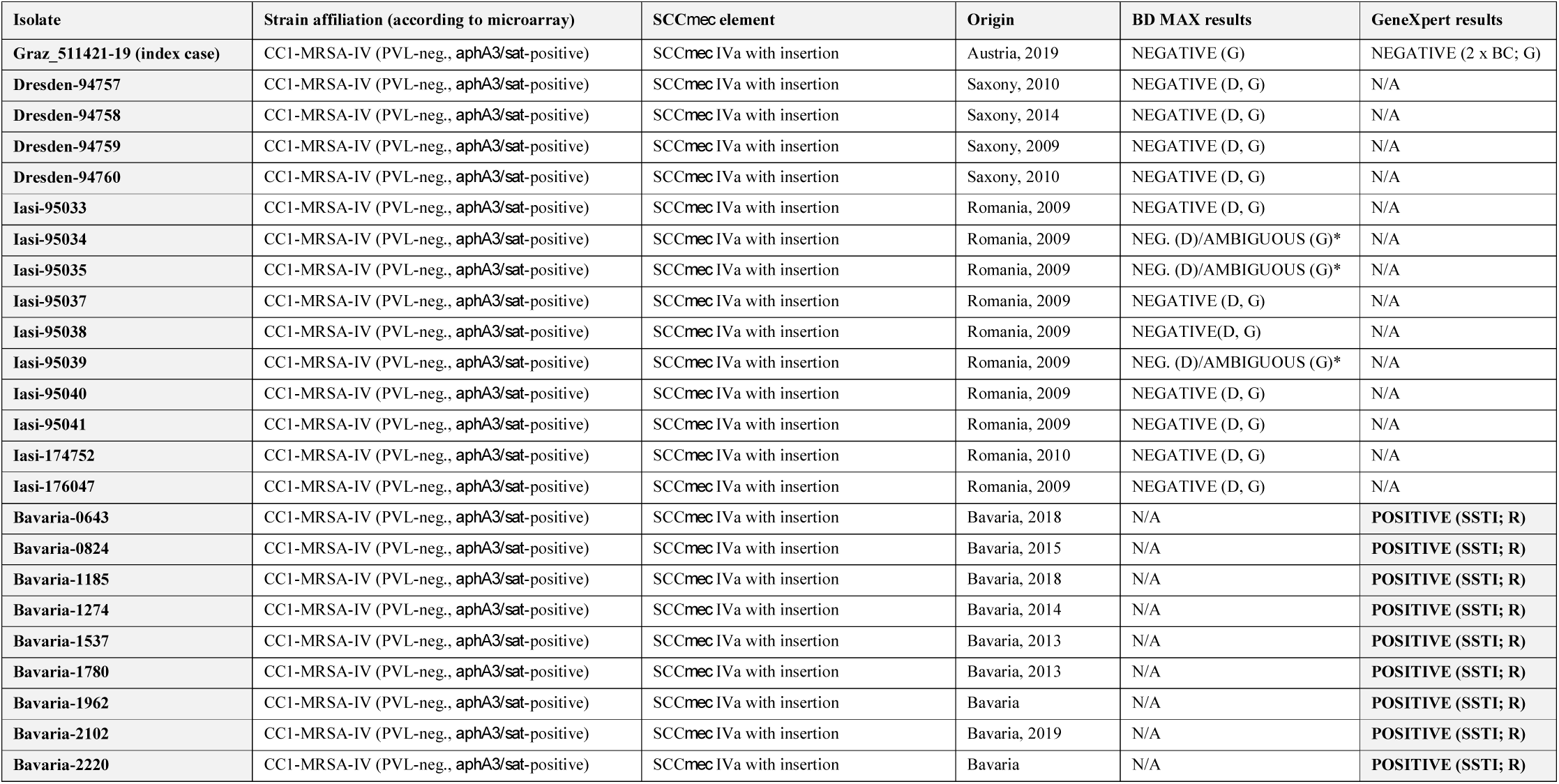
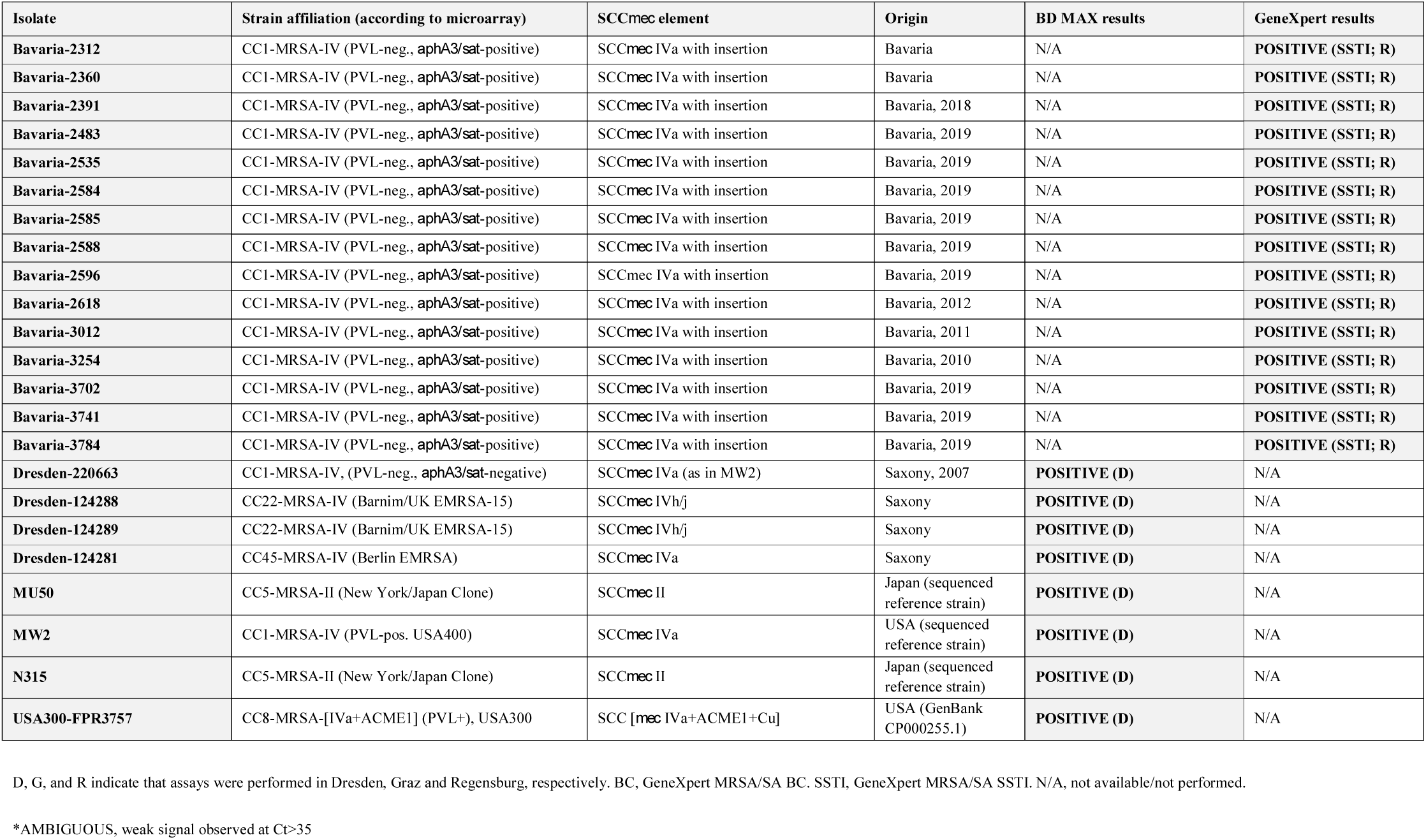
MRSA strains and isolates investigated in the present study and their detection using commercially available *orfX*/SCC*mec* junction site assays.

Nineteen Bavarian isolates recovered at the University Medical Centre in Regensburg between 2010 and 2019 were included, along with five isolates from other tertiary care centres in Bavaria. In Bavaria, the European CC1-MRSA-IV strain appears to be common. At the University Medical Centre in Regensburg, retrospective microarray profiling-based typing of 3,067 isolates revealed a clear increase in the prevalence of this strain. It accounted for <1% of typed MRSA between 2010 and 2013, but reached a prevalence of 9.4% (26 out of 277) in 2019, being the third most common strain (behind CC5-MRSA-II and CC398-MRSA-V/VT) at that centre.

All study isolates were preserved at -80°C in cryovials following identification as MRSA, and molecular typing was performed retrospectively by microarray profiling.

### Cepheid GeneXpert

The index specimen from a positive blood culture was tested with the with the Cepheid GeneXpert system using the MRSA/SA BC assay (Lot. No. 1000148707). It was later re-tested with another lot of the same assay (Lot no. 1000179462). Further investigations on GeneXpert MRSA/SA BC were not possible due to restricted laboratory capacities during the influenza season and the SARS-CoV-2 pandemic.

Twenty-four Bavarian isolates were tested in Regensburg with the Cepheid GeneXpert MRSA/SA SSTI assay (Lot no. 1000180532). It was used according to manufactureŕs instructions starting from pure overnight cultures on CBA.

#### BD MAX

The index isolate was tested in Graz using the BD MAX Staph SR assay (Ref.: 443419; Lot no. 9303156). Additional isolates as well as control strains (see above and Table 1) were grown overnight on Columbia Blood Agar (CBA) and sent for processing to the laboratories in Graz and Dresden where the BD MAX Staph SR Assay (Graz, Lot no. 9303156; Dresden, Lot no. K55928980720210312) was used according to the manufacturer’s instructions.

#### Microarray-based molecular characterisation

Genotyping of all strains was performed using the *S. aureus* Genotyping Kit 2.0 (Abbott [Alere Technologies GmbH]). The microarray-based assay covers 333 different targets related to approximately 170 different genes and their allelic variants. The list of target genes as well as sequences of probes and primers have been described in detail previously along with all relevant protocols (18).

Briefly, *S. aureus* was cultivated and harvested on CBA. Subsequently, DNA extraction was performed with an enzymatic combination of lysis enzymes and buffer from the *S. aureus* Genotyping Kit 2.0 kit and Qiagen DNA extraction columns (Qiagen, Hilden, Germany), according to the manufacturers’ instructions. Linear amplification was performed using one specific primer per target. During amplification, biotin-16-dUTP was incorporated into amplicons. After incubation and several washing steps, hybridisation to the probes on the array was detected using streptavidin horseradish peroxidase that caused local precipitation at the spot where the specific amplicon was bound to the probe. Microarrays were photographed and analysed with a designated reader and software (IconoClust, Abbott (Alere Technologies GmbH)). Array profiling permits rapid isolate characterisation, detects the presence or absence of certain genes or alleles, automatically compares query isolates to a database of existing patterns, and assigns isolates to clonal complexes, known strains and SCC*mec* types.

#### Whole-genome sequencing

All isolates underwent whole-genome sequencing (WGS). Genomic DNA was extracted as detailed above for microarray-based molecular characterisation, and its quality was assessed as previously described (19). The Nextera DNA Flex Library Preparation Kit (Illumina, Eindhoven, The Netherlands) was used according to the manufacturer’s instructions and libraries underwent paired-end sequencing using the 500-cycle MiSeq Reagent Kit v2 (Illumina). Libraries were scaled to exhibit at least 50-fold coverage and the quality of each sequencing run was assured following cluster density and Q30 assessment. Raw sequence reads were trimmed using fastp version 0.19.11 (20). Trimmed sequence read sets underwent *de novo* assembly using SPAdes version v3.9.1 (21) and all contigs under 1,000 bp were removed.

### RESULTS

#### Cepheid GeneXpert tests and BD MAX

Test results are provided in detail in Table 1.

Testing of the index MRSA isolate with two different lots of the GeneXpert MRSA/SA BC assay yielded negative results. However, the GeneXpert MRSA/SA SSTI assay correctly identified 24 European CC1-MRSA-IV isolates as MRSA.

The BD MAX assay system failed to identify isolates (n = 15) of the European CC1-MRSA-IV clone although control reference MRSA strains handled in parallel were correctly identified.

### Description of the European CC1 strain

Microarray profiling and WGS analysis showed that the index MRSA isolate (Graz_511421–19) belonged to a PVL-negative CC1-MRSA-IV clone previously described as “European CC1-MRSA-IV” (13, 14, 17). This clone has been identified in Romania, Ireland and Germany (13, 14, 17) and is represented by a published genome sequence (RBVO00000000.1; (14)).

European CC1-MRSA-IV isolates typically exhibit sequence type (ST) 1 (1–1–1–1–1–1–1) or its single locus variant ST4110 (1–1–1–1–1–1–558), and *spa* type t127 (07–23–21–16–34–33–13) or to related types such as t386 (07–23–13) or t13790 (07–23–21–16–34–33–34–34–33–34).

This clone is resistant to beta-lactam antimicrobials due to a functional *mecA* encoded on an SCC*mec* IV element (see below). It is usually resistant to erythromycin and clindamycin (*ermC*), tetracycline (*tetK*), as well as to kanamycin and neomycin (*aphA*3, accompanied by *aadE* and *sat*). Some isolates of this clone also carry *aacA-aphD* and are resistant to gentamicin. This was not the case for the index isolate from Graz. A particular variant of this clone from Ireland is resistant to mupirocin (*mupR*) and quaternary ammonium compounds such as chlorhexidine due to the presence of a plasmid encoding *iles2/mupR* and *qacA*, as previously described (14). Fusidic acid resistance has not yet been detected in this epidemic clone. In CC1, this property is usually encoded by SCC*mec*-borne *fusC*, which indicates affiliation to other CC1 strains (of Middle Eastern or Southern Hemisphere background); and *fusB/far*-1 was not detected in isolates tested hitherto.

European CC1-MRSA-IV is PVL-negative and only very rarely carries supplementary enterotoxin genes (*sek/seq*) in addition to *seh* that is ubiquitously present in CC1.

### Description of the SCC*mec* element of the European CC1 strain

The SCC*mec* element sequences of the European CC1-MRSA-IV clone were essentially identical in all study isolates as well as in the previously sequenced Irish isolate (GenBank RBVO). Minor polymorphisms were noted within *dru* and within the region between Q2FKL7 and Q8VUV8. These were not relevant for the present study, and can be considered random duplications/deletions, or possibly even as assembly artefacts.

This CC1-MRSA strain has an SCC*mec* IVa element. In contrast to MW2-like SCC*mec* IVa however, it harbours an insertion of ca. 5,350 nucleotides, adjacent to *orfX*. This sequence starts with a novel SCC terminal sequence alternate to *dcs* (“SCCterm 15”; see below) and encodes six different hypothetical proteins (E7MHX1, *ydiL*2, C5QAP8, A8YYX4, *npd*, H4AYD7; nucleotide positions 280,690–286,024 of GenBank RBVO000005.1). Compared to MW2 (BA000033.2), this insertion replaces *dcs* and Q9XB68*-dcs*, and removes the larger part (212 of 240 nt) of the gene encoding hypothetical protein, Q7A213.

The entire gene content of the SCC*mec* IV element is listed in Table 2, and Figure 1 provides a schematic representation of the MGE, including a comparison to MW2 (BA000033.2) and to the sequence of an *S. epidermidis* composite SCC*mec* element (see below).

**Table 2:**
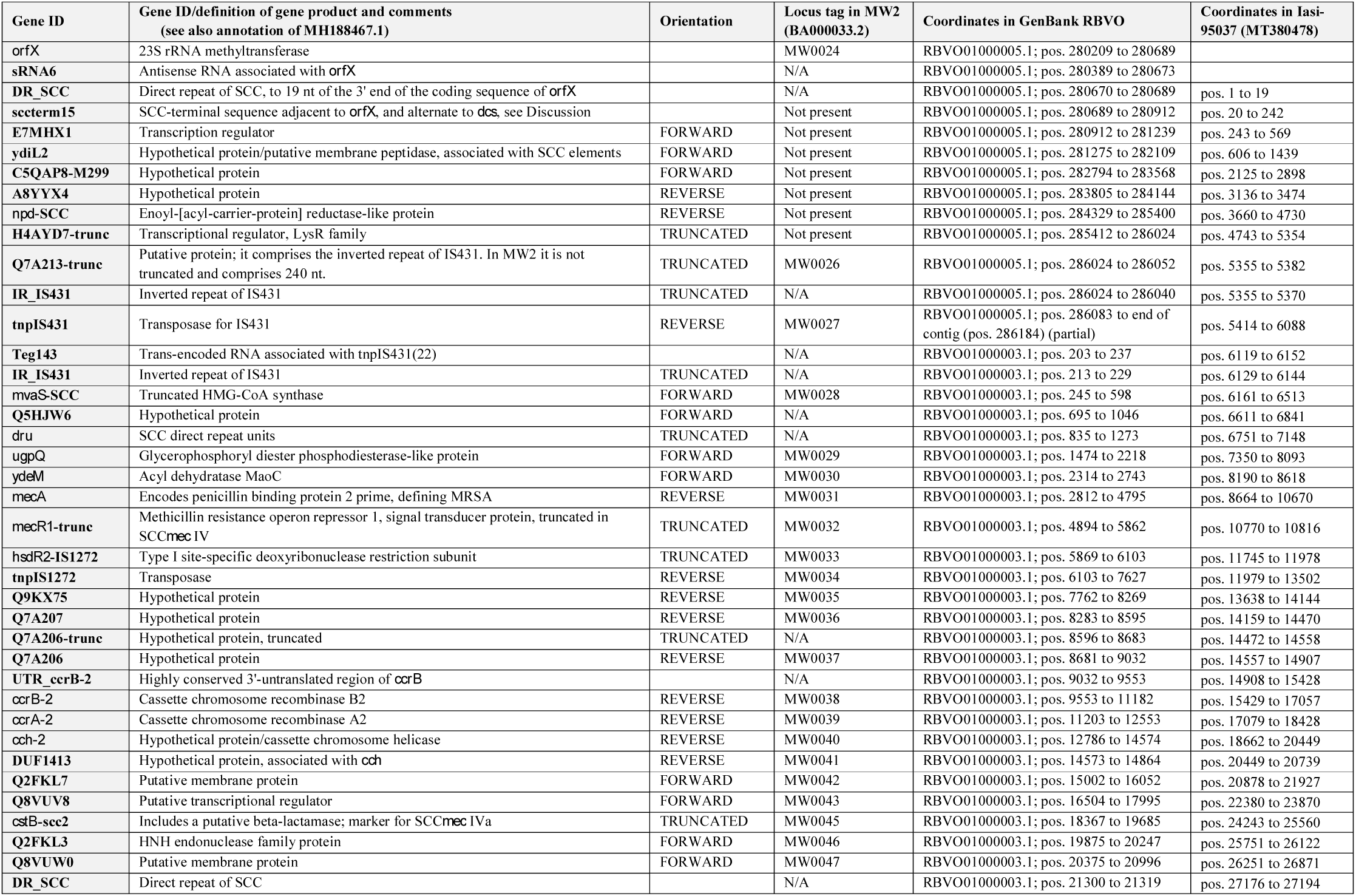
Genes in the variant SCC*mec* IVa element of the European CC1-MRSA-IV strain.

**Figure 1:**
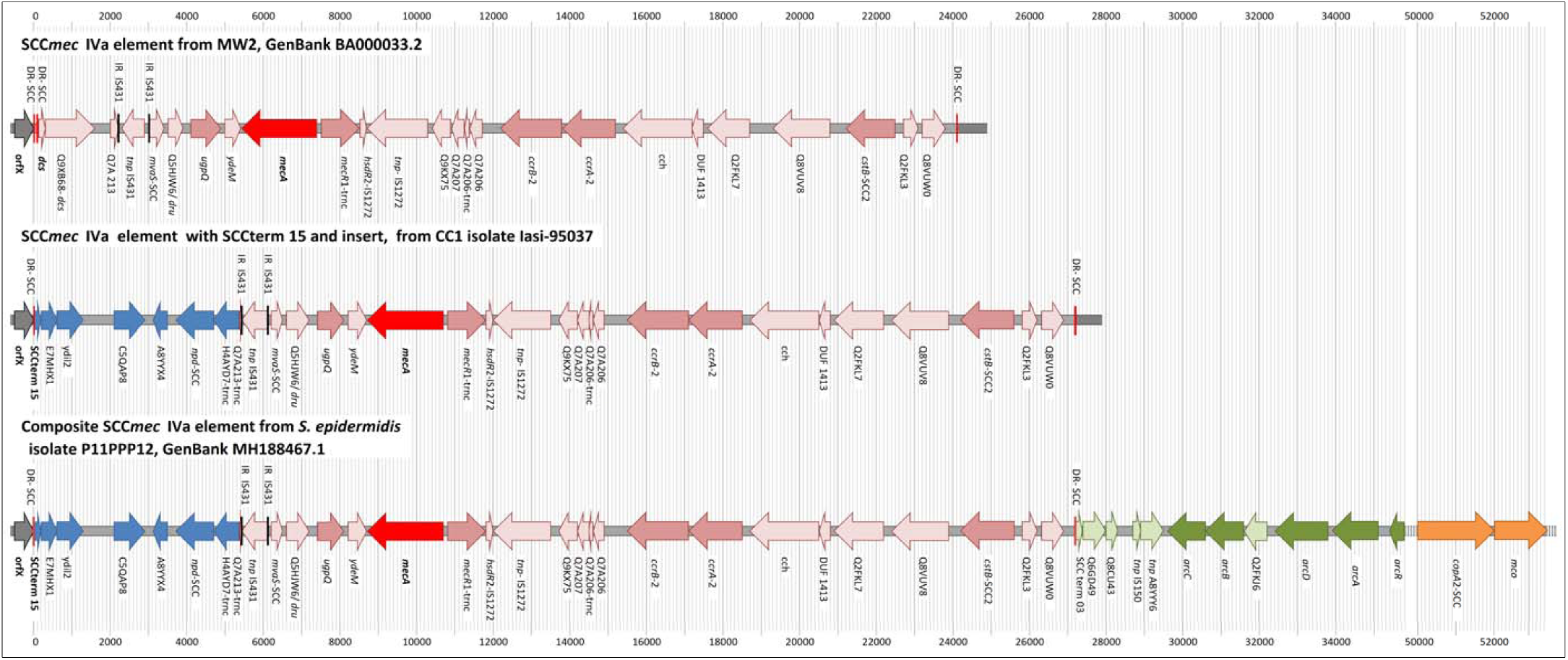
Graphic representation of the SCC*mec* elements in the CC1 reference sequence MW2 (GenBank BA000033.23), in the Euro ropean CC1-MRSA-IV isolate Iasi-95037 (GenBank submission pending) and in the *S. epidermidis* isolate P11PPP12 (GenBank MH188467.1).

## DISCUSSION

The study demonstrates that a CC1-MRSA-IV strain prevalent in Europe can yield false-negative results with frequently used commercial MRSA assays (GenXpert MRSA/SA BC and BD MAX Staph SR). Interestingly, CC1-MRSA-IV strains yielded correctly positive results when using the GeneXpert MRSA/SA SSTI, indicating that the tests are not equipped with the same primers and/or that there is already an updated version for one MRSA test available from Cepheid. Unfortunately, this is not the case for the blood culture (MRSA/SA BC) version that is primarily used for diagnosis in invasive infections. The primers used in these commercial tests are not disclosed, however, the absence of *dcs* and the presence of a large insert in the region downstream of *orfX* correlate with negative GenXpert MRSA/SA BC test and BDmax results. It can therefore be assumed that the diverging results of the assays are related to their coverage of a sequence fragment alternate to *dcs* (SCCterm 15; see Table 2 and Figure 1).

This sequence (SCCterm 15) has previously been observed in at least three different *S. aureus* strains (for discussion of *S. epidermidis*, see below). In CC152-MRSA-XIII, SCCterm 15 is located adjacent to *orfX* (GenBank MG674089, CP024998; (23)) and thus, may affect MSRA assays spanning the *orfX*/SCC*mec* junction. For the remaining two strains, this sequence is situated away from *orfX*, within complex composite SCC*mec* elements. One of these strains is a Danish CC8-MRSA strain (24) with an SCC*mec* IVa/ACME composite element (*e.g*., Strain M1, GenBank HM030720.1). In this strain, the same insert as in the “European CC1” strain is localised at the boundary between an ACME-II element and an SCC*mec* element (25). Here, SCCterm 15 is not adjacent to *orfX*, and the negative junction site PCR results observed can be attributed to yet another SCCterm sequence. Another, CC22-MRSA, strain (CMFT3119, GenBank HF569105.1) from Saudi Arabia also harboured SCCterm 15, E7MHX1, *ydiL*2, IR_IS431 and tnpIS431 localised between a copper resistance element and a composite element encompassing ACME-II and SCC*mec* IVh/j (26).

SCCterm 15, E7MHX1, *ydiL*2, C5QAP8, A8YYX4, *npd* and H4AYD7 are also present in *S. epidermidis* P11PPP12 (GenBank MH188467.1), suggesting a transfer of this region between coagulase-negative staphylococci and CC1 *S. aureus*. Beyond these genes, the entire SCC*mec* IV element of P11PPP12 is identical to that in the CC1-MRSA strain. Most significantly, the fault line cutting short Q7A213 is conserved in both strains (position 5818/5819 in Supplemental file S1), suggesting it unlikely that the insert alone was transferred from a P11PP12-like ancestor into *S. aureus* CC1. Indeed, it is more likely that the *entire* variant SCC*mec* IVa cassette (including the insert) was transferred between the ancestors of the two strains, although the direction of this transfer remains unknown. P11PPP12 also harbours an ACME-II element downstream of SCC*mec* IVa, which comprises another SCC terminal sequence (as in FR753166.1, pos. 481–568), *arc* genes, another copy of tnpIS431, and heavy metal resistance genes. ACME-II is absent from the European CC1-MRSA-IV clone. Therefore, either it must have been lost during or after transfer of the SCC element between *S. epidermidis* and CC1, or it was later acquired by ancestors of *S. epidermidis* P11PPP12. Most likely, the MRSA emerged following transfer of an “atypical” (*dcs*-negative) SCC*mec* IVa element from coagulase-negative staphylococci into a CC1-MSSA strain that is prevalent in South-Eastern Europe.

The presence of MRSA that are negative in *orfX*/SCC*mec* junction site PCRs provides reasons for concern. Firstly, molecular assays are used to predict a presence of MRSA in positive blood cultures and, hence, to prompt administration antibiotics that cover MRSA. A false negative result would harm the patient by delaying effective therapy until conventional susceptibility test results become available. This could easily have fatal consequences. The use of molecular assays for the guidance of therapy in life-threatening circumstances is beneficial for a vast majority of patients due to speed. However, these assays can only detect target sequences that were considered upon primer design. Conventional susceptibility tests are slower but they are not constrained by the choice of primers or by the presence of unknown or unexpected genotypes. Thus, they must not be neglected and should be carried out in parallel to molecular assays.

Secondly, molecular assays are used to guide IPC. A false negative result may result in failure to isolate an infected patient and further spread of the strain. Regarding IPC, it is also noteworthy that this strain can harbour plasmid-borne genes *qacA* and *ileS2/mupR* (14), encoding resistance to quaternary ammonium compounds, biguanides and mupirocin, with the latter compounds commonly being recommended for MRSA decolonisation (27). Thus, it may not only escape detection by molecular screening, but also circumvent decolonisation.

A third and less obvious consequence is a potential shift in the clonal structure of local MRSA population with molecular assays themselves providing a selective pressure favouring the false-negative strain. Other strains will get “penalised” for being PCR-positive as subsequent interventions would hinder their proliferation and transmission. In the long term, this would lead to a higher prevalence of the false-negative strain, and increasingly more therapy and IPC failures.

As previously discussed, the European CC1-MRSA-IV clone may have emerged in South-Eastern Europe. A putative methicillin-susceptible ancestor is common in Romania where the MRSA strain was early and frequently observed (14, 17). Significant case numbers have been reported from Bavaria (14), North Rhine-Westphalia (28), Italy (29) and Ireland (14). Sporadic cases have been described in Austria (30) and Saxony (11).

A strategy to contain the CC1 strain must rely on conventional susceptibility tests or updated molecular tests. We therefore propose to screen, at least, patients with conspicuous travel histories as well as medical or nursing staff recruited in epidemic regions, not only by molecular means but also by culture of nasal swabs on selective growth media. Sequence information and isolates described herein can be used by test manufacturers to adapt future versions of their assays in order to identify this strain.

The problem of unknown or unexpected genotypes also represents a considerable economic risk for the manufacturer of molecular point-of-care tests. The staging and approval of CE–marked in vitro diagnostic tests is very expensive and time-consuming, while there is an absolute necessity to adapt tests to constant evolution of targeted pathogens. It is therefore important for the manufacturers to be able to integrate new markers or new sequence variants into existing tests and platforms with minimal regulatory and economic effort. Unfortunately, the European Commission’s new In Vitro Diagnostic Regulation (IVDR 2017/746) will further raise the regulatory burden (from 2022 on), which may discourage companies from frequently updating assays. Thus, this regulation might, paradoxically, endanger the patients it was intended to protect.

Finally, it is crucial to monitor the emergence of new SCC*mec* junction sites in staphylococci, both in *S. aureus* and coagulase-negative staphylococci, as mobile SCC*mec* elements can be readily transmitted between different strains and species, as was the case in the strain described herein. It is imperative that companies that offer rapid tests should react to these changes and adapt their products to the market as quickly as possible.

## Data Availability

Relevant data are either covered by the manuscript and the supplemental file, or (in case of one sequence) are suvbmitted to GenBank; accession no. MT380478, currently still being processed)

## AUTHORŚ CONTRIBUTIONS

**Stefan Monecke**: Conception and design of the study, data visualisation, analysis and interpretation of data, drafting of the manuscript

**Elisabeth König:** Collection of strains and data generation, analysis and interpretation of data

**Megan R. Earls:** Data generation, analysis and interpretation of data; drafting of the manuscript

**Eva Leitner:** Conception and design of the study, data generation, analysis and interpretation of data, drafting of the manuscript

**Elke Müller:** Data generation, analysis and interpretation of data

**Gabriel Wagner:** Data generation, analysis and interpretation of data

**David Poitz:** Data generation, analysis and interpretation of data

**Lutz Jatzwauk:** Data generation, analysis and interpretation of data

**Teodora Vremerǎ:** Collection of strains and data generation

**Olivia S. Dorneanu:** Collection of strains and data generation

**Alexandra Simbeck:** Collection of strains and data generation, analysis and interpretation of data

**Andreas Ambrosch:** Data generation, analysis and interpretation of data

**Ines Zollner-Schwetz:** Data generation, analysis and interpretation of data

**Robert Krause:** Data generation, analysis and interpretation of data

**Werner Ruppitsch:** Data generation, analysis and interpretation of data

**Wulf Schneider:** Data generation, analysis and interpretation of data

**David C. Coleman:** Analysis and interpretation of data; drafting of the manuscript

**Ivo Steinmetz:** Analysis and interpretation of data; drafting of the manuscript

**Ralf Ehricht:** Conception and design of the study, analysis and interpretation of data; drafting of the manuscript

## ACKNOWLEDGMENTS

DC and ME wish to acknowledge the support of the staff of the Irish National MRSA Reference Laboratory at St. James’s Hospital, Dublin, Ireland.

## COMPETING INTEREST

Not applicable.

## ETHICS APPROVAL AND CONSENT TO PARTICIPATE

Not applicable.

## FUNDING

There was no external funding for this study.

## SUPPLEMENTS

**Supplemental File S1:** Alignment of the SCC*mec* elements of study isolates, reference strains and the individual SCC-associated genes.

**Supplemental File S2:** Sequence of the SCC*mec* element of study isolate Iasi-95037 (GenBank MT380478, submission being processed)

## Notes

### Competing Interest Statement

The authors have declared no competing interest.

